# Structural Elucidation of β_1_- and β_2_-Transferrin Using Microprobe-Capture In-Emitter Elution and High-Resolution Mass Spectrometry

**DOI:** 10.1101/2023.01.29.23285161

**Authors:** Ruben Yiqi Luo, Christopher Pfaffroth, Samuel Yang, Kevin Hoang, Priscilla S.-W. Yeung, James L. Zehnder, Run-Zhang Shi

## Abstract

**Background:** Cerebrospinal fluid (CSF) leak is typically diagnosed by detecting a protein marker β_2_-transferrin (β_2_-Tf) in secretion samples. β_2_-Tf and β_1_-transferrin (β_1_-Tf) are glycoforms of human transferrin (Tf). A novel affinity capture technique for sample preparation, called microprobe-capture in-emitter elution (MPIE), was incorporated with high-resolution mass spectrometry (HR-MS) to analyze the Tf glycoforms and elucidate the structures of β_1_-Tf and β_2_-Tf.

**Methods:** To implement MPIE, an analyte is first captured on the surface of a microprobe, and subsequently eluted from the microprobe inside an electrospray emitter. The capture process is monitored in real-time via next-generation biolayer interferometry (BLI). When electrospray is established from the emitter to a mass spectrometer, the analyte is immediately ionized via electrospray ionization (ESI) for HR-MS analysis. Serum, CSF, and secretion samples were analyzed using MPIE-ESI-MS.

**Results:** Based on the MPIE-ESI-MS results, the structures of β_1_-Tf and β_2_-Tf were solved. As Tf glycoforms, β_1_-Tf and β_2_-Tf share the amino acid sequence but have varying N-glycans. β_1_-Tf, the major serum-type Tf, has two G2S2 N-glycans on Asn413 and Asn611. β_2_-Tf, the major brain-type Tf, has an M5 N-glycan on Asn413 and a G0FB N-glycan on Asn611.

**Conclusions:** The structures of β_1_-Tf and β_2_-Tf were successfully elucidated by MPIE-ESI-MS analysis. The resolving power of the novel MPIE-ESI-MS method was demonstrated in this study. On the other hand, knowing the N-glycan structures on β_2_-Tf allows for the design of other novel test methods for β_2_-Tf in the future.

## Introduction

Cerebrospinal fluid (CSF) leak can occur as a result of laceration, blunt trauma, or surgery. It may lead to potentially life-threatening meningitis if left untreated (1,2). Because β_2_-transferrin (β_2_-Tf), a proteoform of human transferrin (Tf), is mainly present in CSF and barely detectable in other body fluids (3,4), CSF leak can be diagnosed by detecting β_2_-Tf in any body fluid – most commonly in rhinorrhea or otorrhea secretion samples. The clinical utility and diagnostic value of β_2_-Tf in CSF leak have been demonstrated (5,6). β_2_-Tf, together with the typical Tf proteoform in serum β_1_-transferrin (β_1_-Tf), were named after their electrophoretic mobility in gel electrophoresis (7). However, the structures of β_1_-Tf and β_2_-Tf have not been elucidated.

There has been extensive basic research on human Tf since it is a high-abundance protein in human blood with a major role in iron metabolism. The amino acid sequence of Tf precursor was determined through protein cleavage and cDNA characterization, showing a full sequence of 698 amino acids; after the removal of an N-terminal signal peptide, the mature form of Tf contains 679 amino acids, 19 intramolecular disulfide bonds formed between cysteine residues, and two N-glycans attached to the amino groups of the side chains of Asn419 and Asn611 (Asn432 and Asn630 of Tf precursor) (8–11). Tf proteoforms typically vary by the N-glycan structures (11– 13), and the Tf proteoforms of interest so far are Tf glycoforms, including β_1_-Tf and β_2_-Tf (7).

Although it is known that β_2_-Tf has desialylated N-glycans while β_1_-Tf has fully sialylated N-glycans (3,14,15), the structures of β_1_-Tf and β_2_-Tf have not been clarified. On the other hand, the N-glycans on Tf glycoforms in serum and in CSF were characterized using gel electrophoresis, liquid chromatography, and enzymatic digestion-based mass spectrometry (MS) by neurobiologists (13,16–19). It was reported that a group of Tf glycoforms were present in serum, namely serum-type Tf or sTf, among which a specific Tf glycoform (major serum-type Tf) predominated (12,13); the serum-type Tf also existed in CSF, and the major serum-type Tf used to be named Tf-2 in CSF (18); an additional group of Tf glycoforms were present in CSF, namely brain-type Tf or Tf-1 in CSF, among which a specific Tf glycoform (major brain-type Tf) was more abundant than the rest (15,20). The brain-type Tf was at least partly synthesized in the CSF-producing tissue choroid plexus rather than being produced by glycosidase digestion of serum-type Tf (16,20). It was found that the N-glycans on the major serum-type Tf consisted of bi-antennary oligosaccharide chains with sialylated terminals, and those on the brain-type Tf are desialylated, or more specifically, unsialylated (asialotransferrin) (15,16,18,20). It was hypothesized that the major brain-type Tf was β_2_-Tf (21), but this hypothesis has not been proved.

In clinical cases where CSF leak is suspected, secretion samples can be collected from patients and sent to clinical laboratories to test for the presence of β_2_-Tf. The conventional method to test β_2_-Tf as well as β_1_-Tf is agarose gel immunofixation electrophoresis (IFE) (2–4). Although it is widely used in clinical laboratories, it does not provide structural information of the analytes. Thus, the structures of β_1_-Tf and β_2_-Tf, particularly the N-glycan structures on these Tf glycoforms, remained an unanswered question with this method.

As an emerging technology in clinical diagnostics, high-resolution mass spectrometry (HR-MS), particularly top-down HR-MS, can be used to analyze a protein target in its intact state and elucidate the post-translational modifications and amino acid variations in its proteoforms (22– 24). While HR-MS is an ideal tool to analyze the Tf glycoforms, the quality of data acquired during HR-MS analysis depends on sample preparation (25,26). In this article, a novel affinity capture technique for sample preparation, called microprobe-capture in-emitter elution (MPIE), was incorporated with HR-MS to study the Tf glycoforms (27). MPIE can directly couple a label-free optical sensing technology with MS. The label-free optical sensing technology is next-generation biolayer interferometry (BLI, also named as thin-layer interferometry, TFI), which senses optical thickness changes on the sensing surface of a microprobe caused by biomolecular interactions, achieving real-time measurement without employing a reporter molecule (enzyme, fluorophore, etc.) (28,29). To implement MPIE, an analyte is first captured on the surface of a microprobe, and subsequently eluted from the microprobe inside an electrospray emitter. The capture process is monitored in real-time via BLI. When electrospray is established from the emitter to a mass spectrometer, the analyte is immediately ionized via electrospray ionization (ESI) for HR-MS analysis. By this means, BLI and HR-MS are directly coupled in the form of MPIE-ESI-MS, which is readily deployed to analyze the Tf glycoforms and elucidate the structures of β_1_-Tf and β_2_-Tf. The study can pave a way for the development of novel clinical assays for β_2_-Tf.

## Materials and Methods

### Materials and Specimens

LC-MS grade water, acetonitrile, formic acid, and 0.2 µm PVDF syringe filters were purchased from Thermo Fisher Scientific (Waltham, MA). Tf standard (purified sTf) was purchased from Sigma-Aldrich (St. Louis, MI). A mouse monoclonal anti-transferrin IgG antibody (anti-Tf Ab) was obtained from Sinobiological (Wayne, PA), and biotinylated using an EZ-Link HPDP-Biotin reagent kit (Waltham, MA). Remnant CSF and serum samples from patients, and secretion samples from patients suspected of CSF leak were obtained from Stanford Health Care and Stanford Children’s Health, following approved institutional review board protocols for the use of remnant patient specimens.

### Sample Preparation

The biotinylated anti-Tf Ab was diluted in phosphate-buffered saline at pH 7.4 with 0.02% Tween 20 and 0.2% BSA (PBST-B) to 10 μg/ml for use. A pooled CSF sample was made by mixing 9 CSF samples from patients to explore the analytical sensitivity of MPIE-ESI-MS for β_2_-Tf in CSF. The pooled CSF sample was mixed with water to make a dilution series. All CSF samples were 1:1 diluted in phosphate-buffered saline at pH 7.4 with 0.02% Tween 20 (PBST) and all serum samples were 1:19 diluted in PBST-B. Secretion samples from patients were first mixed with an equal amount of water and filtered using a 0.2 µm PVDF syringe filter, and then 1:1 diluted in PBST.

To study the gel electrophoresis-separated Tf glycoforms, gel electrophoresis of CSF samples was carried out using a Hydragel 6 β_2_ Transferrin kit (Sebia, Lisses, France) following the manufacturer’s protocol. In brief, a CSF sample was first 1:1 mixed with an iron-saturating solution, and then 10 µl of the sample was loaded to each of the 6 wells on an agarose gel. Gel electrophoresis was implemented in a Hydrasys 2 instrument (Sebia, Lisses, France), and the agarose gel was removed from the instrument after the gel electrophoresis was completed, without running the immunofixation steps. The agarose gel was placed on a paper template marked with the β_1_-Tf and β_2_-Tf band regions, and the gel stripes of the band regions were cut out using a scalpel. Each gel stripe was placed in a 1.5 ml sample tube, 200 µl PBST was added, and the sample was rocked for 2 hr at room temperature to extract the analyte from the gel stripe. After extraction, the supernatant was filtered using a 0.2 µm PVDF syringe filter.

### MPIE-ESI-MS Instrumentation and Experiment

An MPIE-ESI-MS experiment consists of two parts: BLI-based affinity capture and in-emitter elution ESI-MS, with the details including instrumentation described elsewhere (27). The BLI-based affinity capture was implemented in a Gator Plus analyzer (Gator Bio, Palo Alto, CA): a BLI microprobe pre-coated with streptavidin was first dipped into the biotinylated anti-Tf Ab solution for 10 min to load the anti-Tf Ab, then dipped into a sample for 10 min to capture Tf molecules, and rinsed in PBST for 1 min to remove non-specifically bound molecules. The in-emitter elution ESI-MS was implemented in an EMASS-II ESI ion source which coupled an ECE-001 capillary electrophoresis instrument (CMP Scientific, Brooklyn, NY) with an Orbitrap Q-Exactive Plus mass spectrometer (Thermo Scientific, San Jose, CA): an electrospray emitter with a regular open end and a tapered open end (tip orifice diameter 20-30 μm) was filled with a sheath liquid (10 mM ammonium formate in water); after affinity capture, the microprobe was rinsed in the sheath liquid for 10 s, inserted into the emitter through the regular open end, and settled in the tapered end by gravity; the emitter was mounted to the ESI ion source, and a capillary was inserted into the emitter through the regular open end and positioned right behind the microprobe to deliver an elution liquid (80% acetonitrile and 2% formic acid in water); once electrospray was established by applying a positive voltage to the sheath liquid in the emitter, HR-MS data acquisition was initiated, and injection of the elution liquid was started subsequently. The emitter was placed ∼2 mm away from the mass spectrometer inlet with the electrospray voltage set at 2.2 kV. The injection of the elution liquid was driven by 5 psi pneumatic pressure. The following MS parameters were used: ion-transfer capillary temperature 350°C, S-lens RF level 50, and number of microscans 10. Primary mass spectra were acquired in positive polarity at resolution 17.5K.

### Data Analysis

In HR-MS analysis of proteins, it is necessary to deconvolute raw MS data to merge the multiple charge states and isotopic peaks of an analyte to obtain its accurate molecular mass. The acquired data in each MPIE-ESI-MS experiment was viewed as a time trace of MS responses, and the elution time window of an analyte was identified by checking the molecular ions of the analyte at each time point. The data in the elution time window were selected for deconvolution using Biopharma Finder 4.1 (Thermo Fisher Scientific, San Jose, CA) with the ReSpect algorithm. MS peaks of analytes were displayed in deconvoluted mass spectra at uncharged state showing average molecular masses.

## Results

The performance of MPIE-ESI-MS for Tf analysis was demonstrated and reported elsewhere (27). MPIE-ESI-MS had a limit of detection for the Tf standard at 0.063 μg/ml, which was translated to no more than 7 fmol Tf molecules captured on a microprobe (27). The high analytical sensitivity and specificity provided by affinity capture made MPIE-ESI-MS an ideal method to study the Tf molecules in serum, CSF, and secretion samples. The results of a set of samples are shown in Figure 1. The deconvoluted mass spectrum of the serum sample showed a group of MS peaks around 79554 Da; they are mainly serum-type Tf glycoforms and the predominant MS peak at 79554 Da should be the major serum-type Tf (N-glycan structures shown in Figure 1, see details in Discussion). The deconvoluted mass spectrum of the CSF sample showed the major serum-type Tf and a group of MS peaks around 78008 Da; they are mainly brain-type Tf glycoforms and the most abundant MS peak at 78008 Da should be the major brain-type Tf (N-glycan structures shown in Figure 1, see details in Discussion). The deconvoluted mass spectrum of the secretion sample showed both serum-type Tf and brain-type Tf glycoforms, meaning that CSF was present in the sample. This finding was consistent with the fact that the secretion sample was obtained from a patient diagnosed of CSF leak. In addition, the BLI sensorgram (a time trace of label-free optical sensing responses) of each affinity capture process (capture of Tf by anti-Tf Ab on a microprobe) is shown in Figure S1, allowing for realtime monitoring of the capture process.

**Figure 1.**
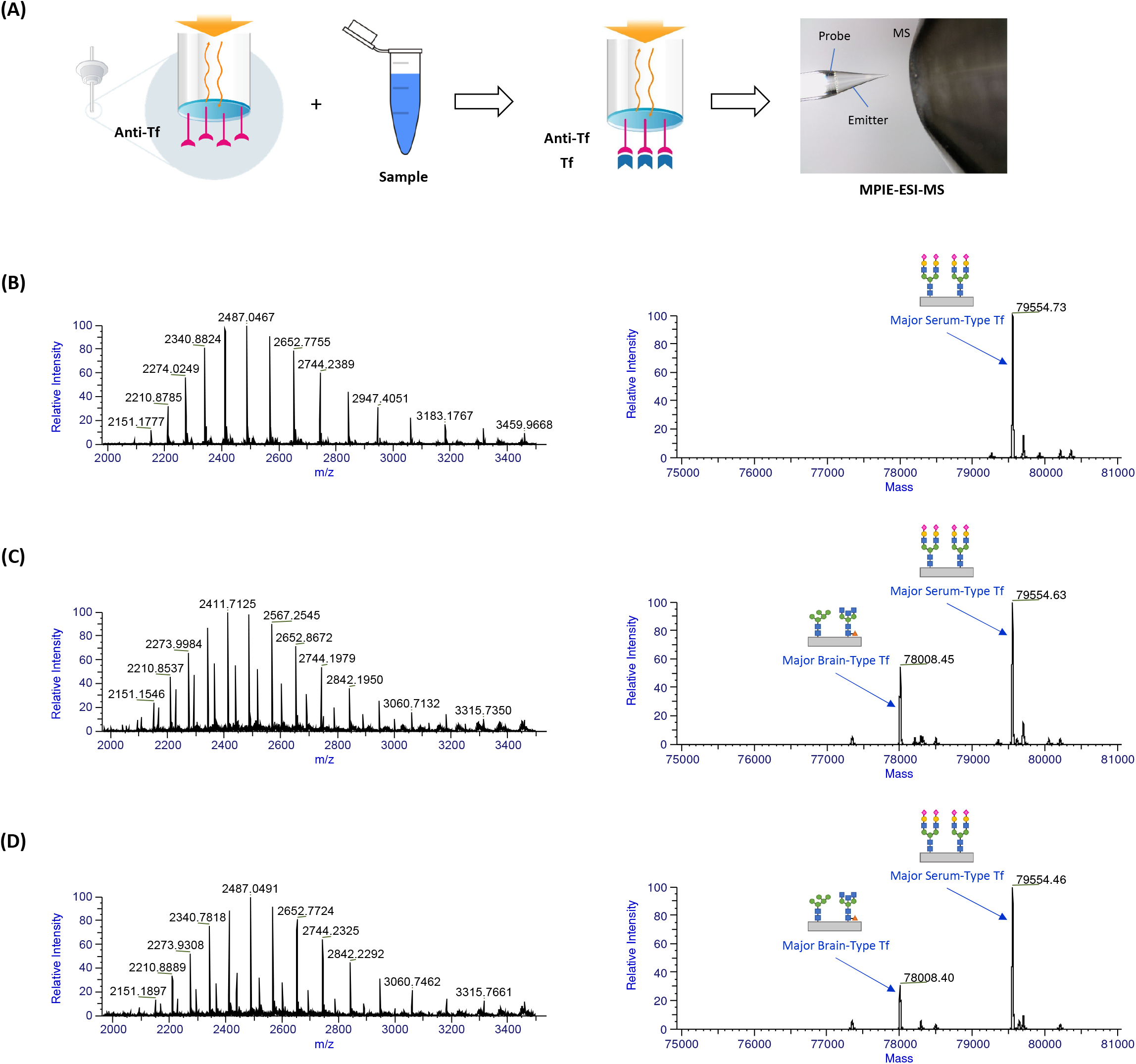
(A) Experiment workflow of MPIE-ESI-MS for Tf analysis. The MPIE-ESI-MS results of (B) a serum sample, (C) a CSF sample, and (D) a secretion sample from a patient diagnosed of CSF leak: HR-MS raw mass spectra (left) and deconvoluted mass spectra (right) of captured Tf molecules, showing serum-type Tf in (B), and both serum-type Tf and brain-type Tf in (C) and (D).

To elucidate the structures of the Tf glycoforms β_1_-Tf and β_2_-Tf, after gel electrophoresis of CSF samples, the extracts from the gel stripes of the β_1_-Tf and β_2_-Tf band regions were analyzed using MPIE-ESI-MS. The results of a set of samples are shown in Figure 2. The deconvoluted mass spectra showed the MS peak of only the major serum-type Tf in the extract from the β_1_-Tf band region and the MS peak of only the major brain-type Tf in the extract from the β_2_-Tf band region. The measured accurate molecular masses were consistent with the experiments in Figure 1 and matched the theoretical molecular masses of the major serum-type Tf and major brain-type Tf (N-glycan structures shown in Figure 2, see details in Discussion). This observation confirmed that β_1_-Tf and β_2_-Tf were actually the major serum-type Tf and major brain-type Tf, respectively. In addition, the gel area between the β_1_-Tf and β_2_-Tf band regions was also analyzed and minor Tf glycoforms in serum and CSF were found, which could be due to the low abundance of the minor Tf glycoforms and limited analyte quantities in a gel stripe.

**Figure 2.**
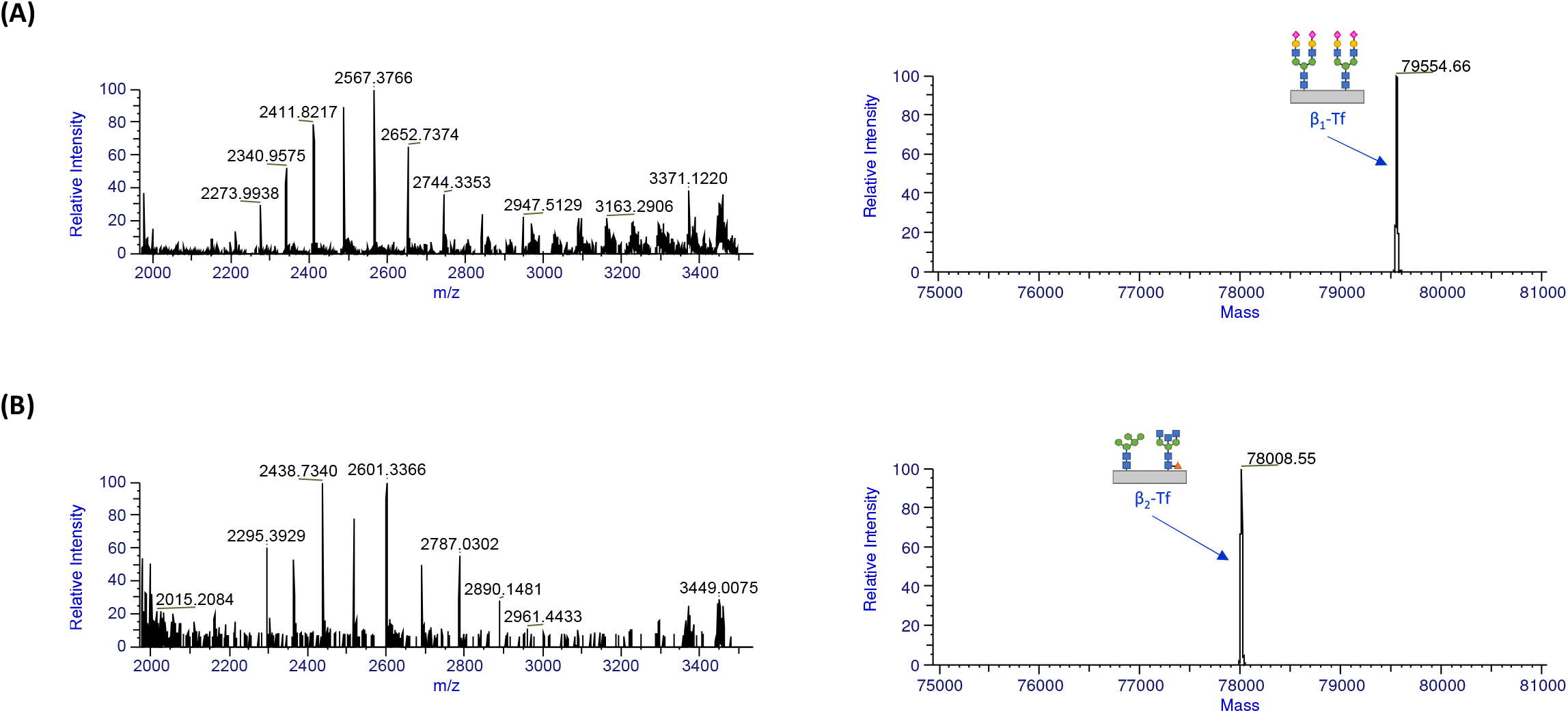
The MPIE-ESI-MS results of (A) the extract from the gel stripe of the β_1_-Tf band region and (B) the extract from the gel stripe of the β_2_-Tf band region: HR-MS raw mass spectra (left) and deconvoluted mass spectra (right) of captured Tf molecules, showing β_1_-Tf in (B) and β_2_-Tf in (C).

A set of 12 secretion samples from patients suspected of CSF leak were analyzed using the MPIE-ESI-MS method, among which 5 samples were positive for β_2_-Tf and the rest were negative as measured by IFE test and clinical manifestations. As shown in Table 1, the MS peak at 78008 Da was observed in the MPIE-ESI-MS results of the 5 positive samples but not found in those of the 7 negative samples, which further verified that β_2_-Tf was indeed the major brain-type Tf. In addition, the analytical sensitivity of the MPIE-ESI-MS method for CSF and secretion samples was explored. A pooled CSF sample was mixed with water at 1:1, 1:4, 1:9, and 1:19 ratios to prepare a dilution series for analysis. Table S1 summarizes the results, listing the MS peak intensities of β_1_-Tf and β_2_-Tf in the deconvoluted mass spectra (the entire time window of Tf elution selected for deconvolution). The peak intensities decreased with the pooled CSF sample dilution, and it was demonstrated that the MPIE-ESI-MS method was able to detect β_2_-Tf in at least 10-fold diluted CSF (1:9 pooled CSF : water mixture).

**Table 1.**
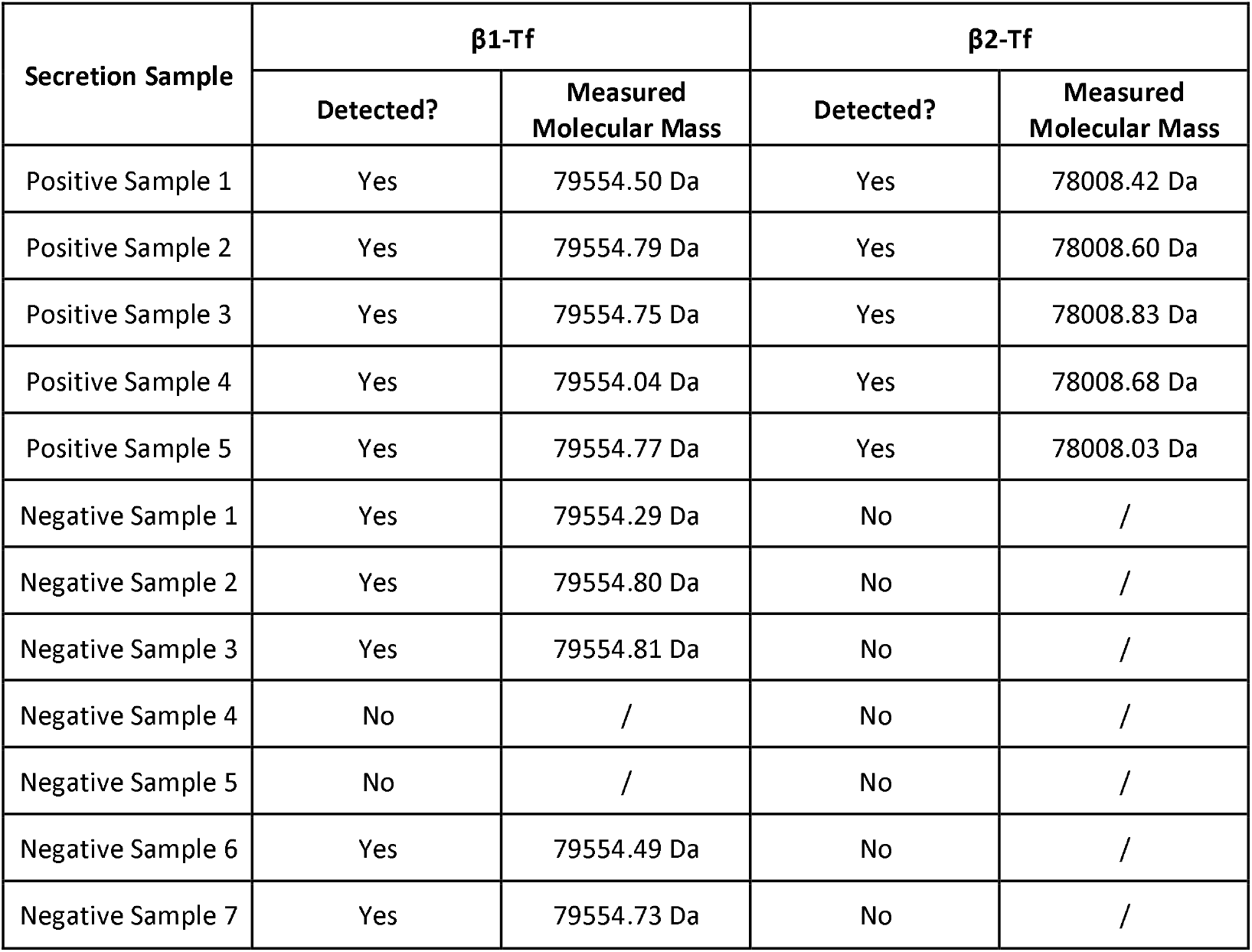
The MPIE-ESI-MS results of 12 secretion samples from patients suspected of CSF leak.

## Discussion

The amino acid sequence of Tf was previously reported (8,9) and the primary structure of a mature Tf molecule is illustrated in Figure 3A. As Tf glycoforms, β_1_-Tf and β_2_-Tf share the amino acid backbone but have varying N-glycans. The N-glycans on proteins typically include bi-, tri-, or tetra-antennary oligosaccharide chains, resulted from sequential action of glycosyltransferases (30,31). An N-glycan can be named according to its sugar composition and branching structure using a few nomenclature systems (traditional nomenclature used in this article) (32). With designated N-glycan types, the accurate molecular mass of a Tf glycoform can be calculated, and the correctness of the N-glycan types can be confirmed by comparing the theoretical molecular mass of the Tf glycoform to the measured accurate molecular mass.

**Figure 3.**
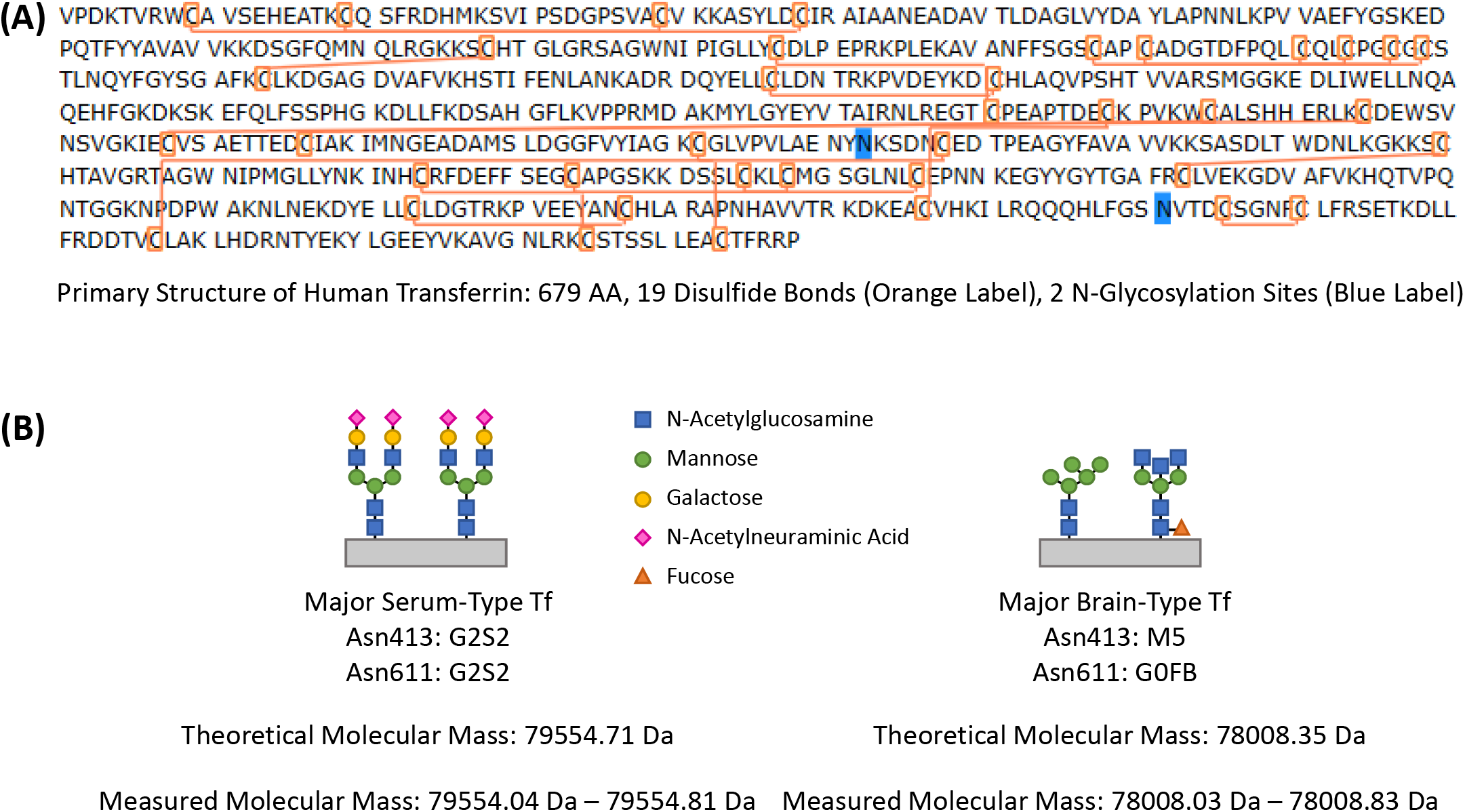
(A) Primary structure of human transferrin, showing the sequence of 679 amino acids, 19 disulfide bonds, and 2 N-glycosylation sites. (B) N-glycan structures on β_1_-Tf (major serum-type Tf) and β_2_-Tf (major brain-type Tf), confirmed by comparing the theoretical molecular masses of the Tf glycoforms with the measured molecular masses in Figure 1, Figure 2, and Table 1.

Regarding the N-glycans on Tf, the previous enzymatic digestion-based MS analysis of the Tf glycoforms in CSF revealed explicitly that the most abundant N-glycan type was G2S2 (17,19). By measuring the trypsin-digested peptides encompassing the glycosylation sites, G2S2 was found consisting of roughly 65% of all N-glycans on Asn413 and 73% of all N-glycans on Asn611 (17). Thus, the Tf glycoform with two G2S2 N-glycans on Asn413 and Asn611 is the major serum-type Tf because (1) it is the most abundant Tf glycoform in serum and CSF, (2) it is fully sialylated (N-acetylneuraminic acid as the sialic acid), and (3) its theoretical molecular mass 79554.71 Da exactly matches the measured molecular mass of the major serum-type Tf (Figure 3B). As the second most abundant Tf glycoform and the most abundant unsialylated Tf glycoform in CSF, the major brain-type Tf should be associated with N-glycan types M5 and G0FB (Figure 3B), which were found as the most abundant unsialylated N-glycans (19).

Particularly, M5 was found consisting of roughly 20% of all N-glycans on Asn413 and G0FB was found consisting of 26% of all N-glycans on Asn611 (17). Thus, the major brain-type Tf has an M5 N-glycan on Asn413 and a G0FB N-glycan on Asn611. Its theoretical molecular mass 78008.35 Da exactly matches the measured molecular mass of the major brain-type Tf. The glycosylation sites of the M5 and G0FB N-glycans were confirmed with another study of trypsin-digested glycopeptides from Tf glycoforms in CSF (18). In addition, the presence of a bisecting N-acetylglucosamine (GlcNAc) in the G0FB N-glycan was proved by the previously reported lectin-binding experiments (16), supporting that G0FB is the more favorable N-glycan type on Asn611 than the other isomeric N-glycan type without a bisecting GlcNAc (19). In addition, the MPIE-ESI-MS analysis of the extracts from the gel bands proved that β_1_-Tf and the major serum-type Tf were identical, so were β_2_-Tf and the major brain-type Tf.

Besides the two major Tf glycoforms in CSF (β_1_-Tf and β_2_-Tf), it is known that other minor Tf glycoforms may also exist in CSF, blood, and other body fluids (4,33). As illustrated in the previous literature, there are a variety of unsialylated N-glycans on Tf molecules, and the sialylated N-glycan G2S2 also has diversity such as the more branched form G3S3 (13,17,34). Thus, a number of fully sialylated, partially sialylated, and unsialylated Tf glycoforms can be formed by the various combinations of the two N-glycans on a Tf molecule. In gel electrophoresis, sialic acids bring negative charges to a Tf molecule under neutral or alkaline pH conditions, influencing its electrophoretic mobility. As such, Tf glycoforms migrate in the order of fully sialylated, disialylated, and unsialylated Tf glycoforms, with regard to the number of sialic acids on the N-glycans. Thus, the minor Tf glycoforms can migrate within or between the β_1_-Tf and β_2_-Tf band regions. For instance, the product insert of the Hydragel 6 β_2_ Transferrin kit states that a band of disialylated Tf glycoforms (disialotransferrin) may exist above the β_2_-Tf band (35). On the other hand, the unsialylated Tf glycoforms besides β_2_-Tf can migrate within the β_2_-Tf band region and interfere with the β_2_-Tf detection. The product insert suggests that the ratio of unsialylated to disialylated bands can be used to confirm the presence of β_2_-Tf. The reason of this practice can be explained as follows: when CSF is present in a sample, the abundance of the major unsialylated Tf glycoform β_2_-Tf significantly exceeds the minor unsialylated and disialylated Tf glycoforms, resulting in a high ratio of the β_2_-Tf band to the disialotransferrin band. This practice in IFE is derived from the lack of molecular structure information in gel electrophoresis. However, when MS is employed, such as in the use of MPIE-ESI-MS, it is unnecessary to compare unsialylated and disialylated Tf glycoforms because β_2_-Tf can be specifically detected by its accurate molecular mass, definitively differentiating it from the other Tf glycoforms with different molecular masses.

The demonstration of MPIE-ESI-MS in detection of β_2_-Tf paved a way to establish a MS-based clinical assay for β_2_-Tf. However, more factors need to be taken into consideration for this purpose. When implementing the accurate molecular mass-based detection of β_2_-Tf, Tf variants resulted from genetic polymorphism should be taken into consideration (11,36–39). In theory, the amino acid mutations in Tf variants affect the molecular masses of β_1_-Tf and β_2_-Tf, but they should not change the molecular mass difference between the two Tf glycoforms (1546 Da), provided the two glycosylation sites are not modified. This hypothesis is supported by the MPIE-ESI-MS results of a few Tf variant-containing CSF samples, as shown in Figure S2, but it needs to be proved with a larger number of Tf variant samples. In addition, the glycoforms in serum-type Tf can vary under specific pathophysiological conditions and β_1_-Tf might not be found in those samples due to aberrant glycosylation (12,34). In such cases the detection and identification of β_2_-Tf can be impacted. These topics are beyond the scope of this article and can be investigated in the future when a MS-based clinical assay for β_2_-Tf is being developed.

## Conclusions

The structures of β_1_-Tf and β_2_-Tf were successfully elucidated by MPIE-ESI-MS analysis. The resolving power of the novel MPIE-ESI-MS method was demonstrated in this study. As an innovative affinity capture technique, MPIE facilitates real-time monitoring of the affinity capture process to overcome the lack of process monitoring in conventional affinity capture techniques (27). On the other hand, knowing the N-glycan structures on β_2_-Tf allows for the design of other novel test methods for β_2_-Tf in the future. For instance, it is possible to employ carbohydrate-binding reagents to selectively capture β_2_-Tf for MS analysis or to establish antibody-lectin Sandwich immunoassays.

## Data Availability

All data produced in the present study are available upon reasonable request to the authors.

## Acknowledgment

The authors thank Gator Bio (Palo Alto, CA) and CMP Scientific (Brooklyn, NY) for kindly providing equipment and consumables, and C. Wong, S. Ferolino, L. Calayag, and R. Rieta at Stanford Health Care for collecting samples for this research.

