## Supplemental Material for "Structural Elucidation of β_1_- and β_2_-Transferrin Using Microprobe-Capture In-Emitter Elution and High-Resolution Mass Spectrometry"

Running Title: Elucidation of $\beta_1$- and $\beta_2$-Transferrin Using MPIE Coupled with HR-MS

Corresponding Author: Ruben Yiqi Luo

Address: 3375 Hillview Ave, Palo Alto, CA 94304

Key Words: $\beta_1$-Transferrin, $\beta_2$-Transferrin, MPIE, HR-MS, N-Glycan

### Figure S1

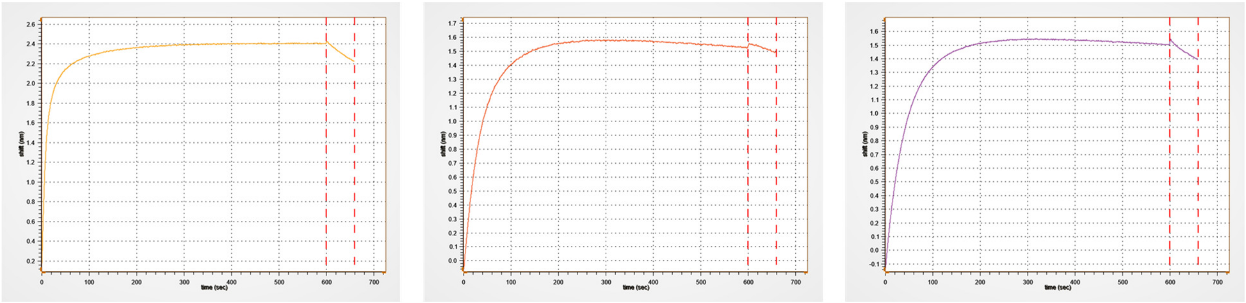

Figure S1. BLI sensorgrams obtained on the 3 microprobes capturing Tf from a serum sample (left), a CSF sample (middle), and a secretion sample from a patient diagnosed of CSF leak (right) (same samples as in Figure 1).

### Figure S2

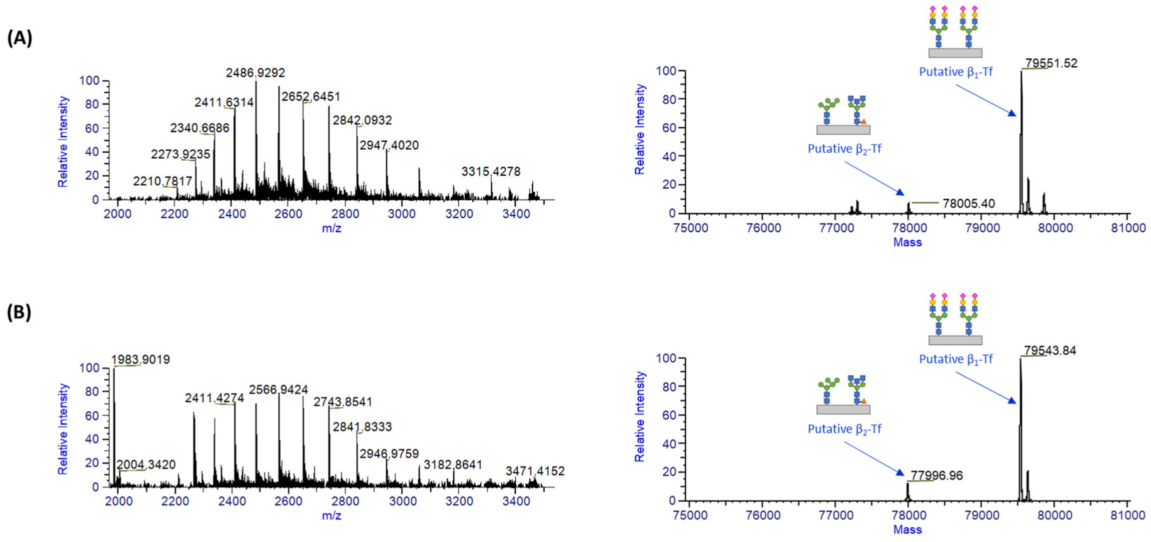


Figure S2. The MPIE-ESI-MS results of two CSF samples containing Tf variants (A) and (B): HR-MS raw mass spectra of captured Tf molecules (left) and deconvoluted mass spectra (right), showing the Tf glycoforms. The molecular masses of the putative $\beta_1$-Tf and $\beta_2$-Tf were different from those of $\beta_1$-Tf and $\beta_2$-Tf in normal CSF samples, but the mass difference between the two Tf glycoforms retained as 1546 Da.

#### Table S1

Table S1. MS peak intensities of $\beta_1$-Tf and $\beta_2$-Tf in the deconvoluted mass spectra (the entire time window of Tf elution selected for deconvolution): a pooled CSF sample was mixed with water at 1:1, 1:4, 1:9, and 1:19 ratios, the neat pooled CSF sample, and the pooled CSF sample spiked with 10 μg/ml and 100 μg/ml Tf standard.

| Sample | MS Peak Intensity of β1-Tf | MS Peak Intensity of β2-Tf |
|---|---|---|
| 1:1 Pooled CSF : Water | $4.10 \times 10^5$ | $1.62 \times 10^5$ |
| 1:4 Pooled CSF : Water | $2.52 \times 10^5$ | $7.03 \times 10^4$ |
| 1:9 Pooled CSF : Water | $9.47 \times 10^4$ | $1.98 \times 10^4$ |
| 1:19 Pooled CSF : Water | $4.18 \times 10^4$ | Not Detected |
